# The Role of Vitamin D in the Age of COVID-19: A Systematic Review and Meta-Analysis

**DOI:** 10.1101/2020.06.05.20123554

**Authors:** Roya Ghasemian, Amir Shamshirian, Keyvan Heydari, Mohammad Malekan, Reza Alizadeh-Navaei, Mohammad Ali Ebrahimzadeh, Hamed Jafarpour, Arash Rezaei Shahmirzadi, Mehrdad Khodabandeh, Benyamin Seyfari, Alireza Motamedzadeh, Ehsan Dadgostar, Marzieh Aalinezhad, Meghdad Sedaghat, Nazanin Razzaghi, Bahman Zarandi, Anahita Asadi, Vahid Yaghoubi Naei, Reza Beheshti, Amirhossein Hessami, Soheil Azizi, Ali Reza Mohseni, Danial Shamshirian

**Affiliations:** Antimicrobial Resistance Research Center, Department of Infectious Diseases, Mazandaran University of Medical Sciences, Sari, Iran.; Department of Medical Laboratory Sciences, Student Research Committee, School of Allied Medical Science, Mazandaran University of Medical Sciences, Sari, Iran.; Gastrointestinal Cancer Research Center, Mazandaran University of Medical Sciences, Sari, Iran.; Student Research Committee, School of Medicine, Mazandaran University of Medical Sciences, Sari, Iran.; Pharmaceutical Sciences Research Center, Department of Medicinal Chemistry, School of Pharmacy, Mazandaran University of Medical Science, Sari, Iran.; Student Research Committee, Golestan University of Medical Sciences, Gorgan, Iran.; Neuromusculoskeletal Research Center, Department of Physical Medicine and Rehabilitation, Iran University of Medical Sciences, Tehran, Iran.; Department of Surgery, Faculty of Medicine, Kashan University of Medical Sciences, Kashan, Iran.; Department of Internal Medicine, Faculty of Medicine, Kashan University of Medical Sciences, Kashan, Iran.; Halal Research Center of IRI, FDA, Tehran, Iran.; Department of Radiology, Isfahan University of Medical Sciences, Isfahan, I.R. Iran.; Department of Internal Medicine, Imam Hossein Hospital, Shahid Beheshti University of Medical Sciences, Tehran, Iran.; Student Research Committee, Iran University of Medical Sciences, Tehran, Iran.; Immunology Research Center, Mashhad University of Medical Sciences, Mashhad, Iran.; Department of Medical Laboratory Sciences, School of Allied Medical Science, Mazandaran University of Medical Sciences, Sari, Iran.; Thalassemia Research Center, Hemoglobinopathy Institute, Mazandaran University of Medical Sciences, Sari, Iran.; Chronic Respiratory Diseases Research Center, National Research Institute of Tuberculosis and Lung Diseases (NRITLD), Shahid Beheshti University of Medical Sciences, Tehran, Iran.

**Keywords:** Pandemic, 2019-nCoV, Coronavirus Outbreaks, SARS-CoV-2, Vitamin D, 25-hydroxyvitamin D, 25(OH)D

## Abstract

**Background:** Evidence recommends that vitamin D might be a crucial supportive agent for the immune system, mainly in cytokine response regulation against COVID-19. Hence, we carried out a systematic review and meta-analysis in order to maximize the use of everything that exists about the role of vitamin D in the COVID-19.

**Methods:** A systematic search was performed in PubMed, Scopus, Embase, and Web of Science up to December 18, 2020. Studies focused on the role of vitamin D in confirmed COVID-19 patients were entered into the systematic review.

**Results:** Twenty-three studies containing 11901participants entered into the meta-analysis. The meta-analysis indicated that 41% of COVID-19 patients were suffering from vitamin D deficiency (95% CI, 29%-55%), and in 42% of patients, levels of vitamin D were insufficient (95% CI, 24%-63%). The serum 25-hydroxyvitamin D concentration was 20.3 ng/mL among all COVID-19 patients (95% CI, 12.1-19.8). The odds of getting infected with SARS-CoV-2 is 3.3 times higher among individuals with vitamin D deficiency (95% CI, 2.5-4.3). The chance of developing severe COVID-19 is about five times higher in patients with vitamin D deficiency (OR: 5.1, 95% CI, 2.6-10.3). There is no significant association between vitamin D status and higher mortality rates (OR: 1.6, 95% CI, 0.5-4.4).

**Conclusion:** This study found that most of the COVID-19 patients were suffering from vitamin D deficiency/insufficiency. Also, there is about three times higher chance of getting infected with SARS-CoV-2 among vitamin D deficient individuals and about 5 times higher probability of developing the severe disease in vitamin D deficient patients. Vitamin D deficiency showed no significant association with mortality rates in this population.

## Introduction

Following the emergence of a novel coronavirus from Wuhan, China, in December 2019, the respiratory syndrome coronavirus 2 (SARS-CoV-2) has affected the whole world and is declared a pandemic by World Health Organization (WHO) on March 26, 2020^1^. According to Worldometer metrics, this novel virus has been responsible for approximately 83,848,186 infections, of which 59,355,654 cases are recovered, and 1,826,530 patients have died worldwide up to January 01, 2021.

After months of medical communities’ efforts, one of the hottest topics is still the role of Vitamin D in the prevention or treatment of COVID-19. Several functions, such as modulating the adaptive immune system and cell-mediated immunity, as well as an increase of antioxidative-related genes expression, have been proven for Vitamin D as an adjuvant in the prevention and treatment of acute respiratory infections ^2–4^. According to available investigations, it seems that such functions lead to cytokine storm suppression and avoid Acute Respiratory Distress Syndrome (ARDS), which has been studied on other pandemics and infectious diseases in recent years ^4–7^.

To the best of our knowledge, unfortunately, after several months, there is no adequate high-quality data on different treatment regimens, which raises questions about gaps in scientific works. On this occasion, when there is an essential need for controlled randomized trials, it is surprising to see only observational studies without a control group or non-randomized controlled studies with retrospective nature covering a small number of patients. The same issue is debatable for 25-hydroxyvitamin D (25(OH)D); hence, concerning all of the limitations and analyze difficulties, we carried out a systematic review and meta-analysis to try for maximizing the use of everything that exists about the role of this vitamin in the COVID-19.

## Methods

### Search Strategy

The Preferred Reporting Items for Systematic Reviews and Meta-Analyses (PRISMA) guideline was considered for the study plan. A systematic search through databases of PubMed, Scopus, Embase, and Web of Science was done up to December 18, 2020. Moreover, to obtain more data, we considered gray literature and references of eligible papers. The search strategy included all MeSH terms and free keywords found for COVID-19, SARS-CoV-2, and Vitamin D. There was no time/location/ language limitation in this search.

### Criteria study selection

Four researchers have screened and selected the papers independently, and the supervisor solved the disagreements. Studies met the following criteria included in the meta-analysis: 1) comparative or non-comparative studies with retrospective or prospective nature; and 2) studies reported the role of vitamin D in confirmed COVID-19 patients. Studies were excluded if they were: 1) *in vitro* studies, experimental studies, reviews, 2) duplicate publications.

### Data extraction & quality assessment

Two researchers (H.J and M.M) have evaluated the papers’ quality assessment and extracted data from selected papers. The supervisor (D.Sh) resolved any disagreements in this step. The data extraction checklist included the name of the first author, publication year, region of study, number of patients, co-morbidity, vitamin D Status, serum 25-hydroxyvitamin D levels, ethnicity, mean age, medication dosage, treatment duration, adverse effects, radiological results, and mortality. The modified Newcastle-Ottawa Scale (NOS) checklist for cross-sectional studies and Jadad scale for clinical trials were used to value the studies concerning various aspects of the methodology and study process.

### Hypothetical strategy

In this case, according to most of the studies, vitamin D cut-off points were considered as follows:

- Vitamin D sufficiency: 25(OH)D concentration greater than 30 ng/mL
- Vitamin D insufficiency: 25(OH)D concentration of 20 to 30 ng/mL
- Vitamin D deficiency: 25(OH)D level less than 20 ng/mL

### Targeted outcomes

1) Frequency of Vitamin D status in COVID-19 patients; 2) Mean 25(OH)D concentration; 3) Association between Vitamin D Deficiency and SARS-CoV-2 infection; 4) Association between Vitamin D Deficiency and COVID-19 severity; 5) Association between Vitamin D Deficiency and COVID-19 mortality; 6) Co-morbidity frequency; 7) Ethnicity frequency.

### Heterogeneity assessment

I-square (*I*^*2*^) statistic was used for heterogeneity evaluation. Following Cochrane Handbook for Systematic Reviews of Interventions ^8^, the *I*^*2*^ was interpreted as follows: “*0% to 40%: might not be important; 30% to 60%: may represent moderate heterogeneity; 50% to 90%: may represent substantial heterogeneity; 75% to 100%: considerable heterogeneity. The importance of the observed value of I^2^ depends on (i) magnitude and direction of effects and (ii) strength of evidence for heterogeneity (e.g., P-value from the chi-squared test, or a confidence interval for I^2^)*.” Thus, the random-effects model was used for pooling the outcomes in case of heterogeneity; otherwise, the inverse variance fixed-effect model was used. Forest plots were presented to visualize the degree of variation between studies.

### Data analysis

Meta-analysis was performed using Comprehensive Meta-Analysis (CMA) *v*. 2.2.064 software. The pooling of effect sizes was done with 95% Confident Interval (CI). The fixed/random-effects model was used according to heterogeneities. In the case of zero frequency, the correction value of 0.1 was used.

### Publication bias & sensitivity analysis

Begg’s and Egger’s tests and the funnel plot were used for publication bias evaluation. A *P*- value of less than 0.05 was considered as statistically significant.

## Results

### Study selection process

The first search through databases resulted in 1382 papers. After removing duplicated papers and first step screening based on title and abstract, 121 papers were assessed for eligibility. Finally, 23 articles were entered into the meta-analysis. PRISMA flow diagram for the study selection process is presented in Figure 1.

**Figure 1.**
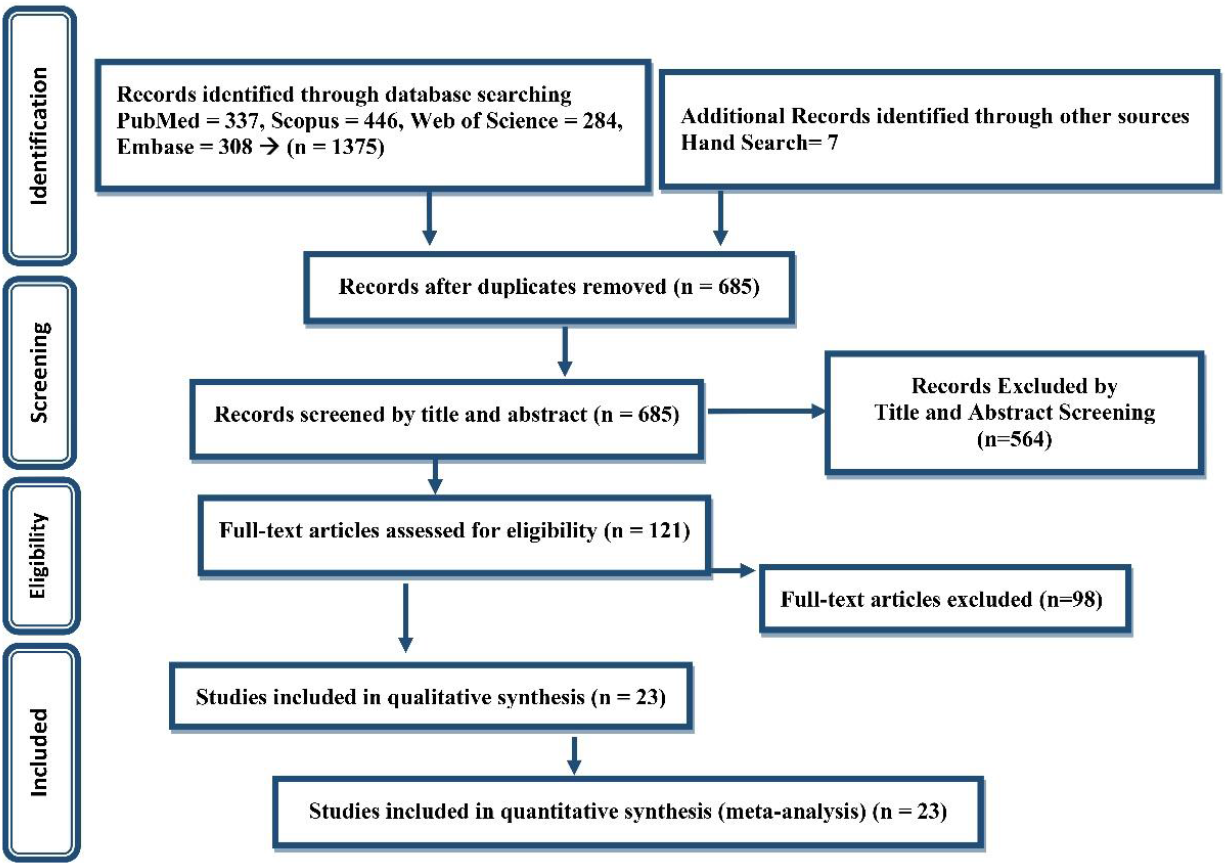
PRISMA flow diagram for the study selection process.

### Study characteristics

Among the 23 studies included in the meta-analysis, all were designed in retrospective nature, except for five study in prospective nature. The studies’ sample size ranged from 19 to 7807, including 11901 participants. Characteristics of studies entered into the systematic review presented in Table 1.

**Table 1.**
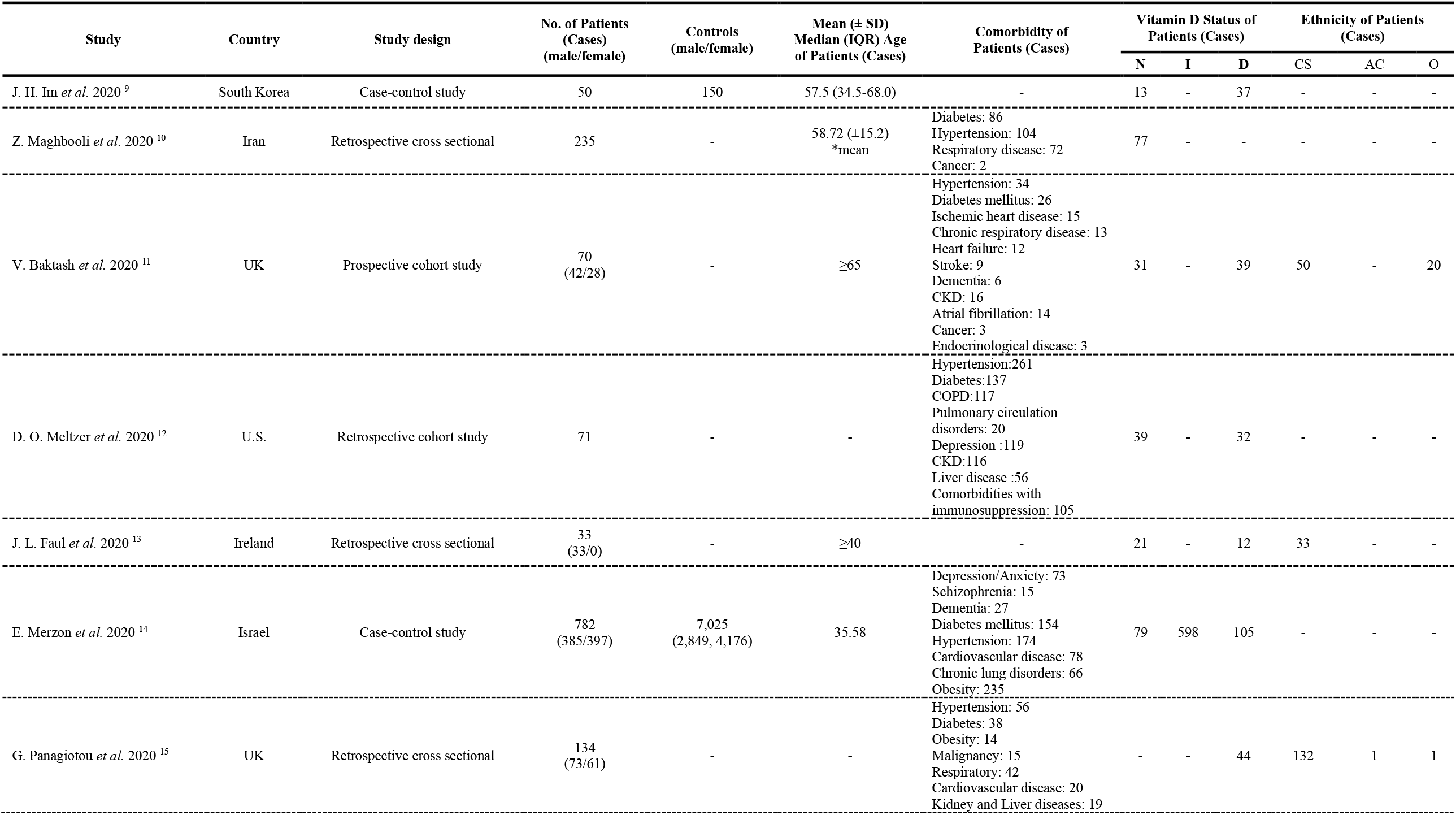
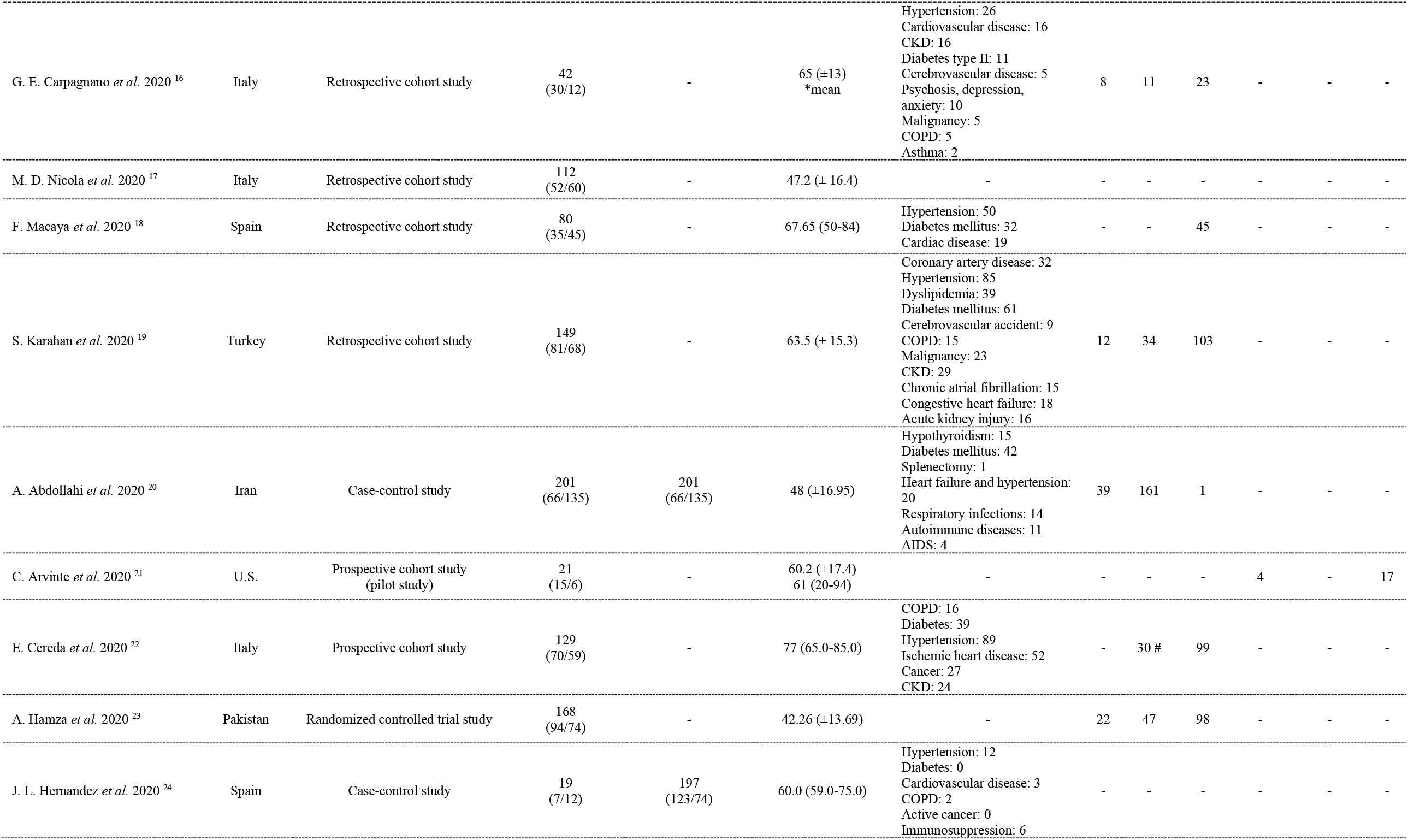
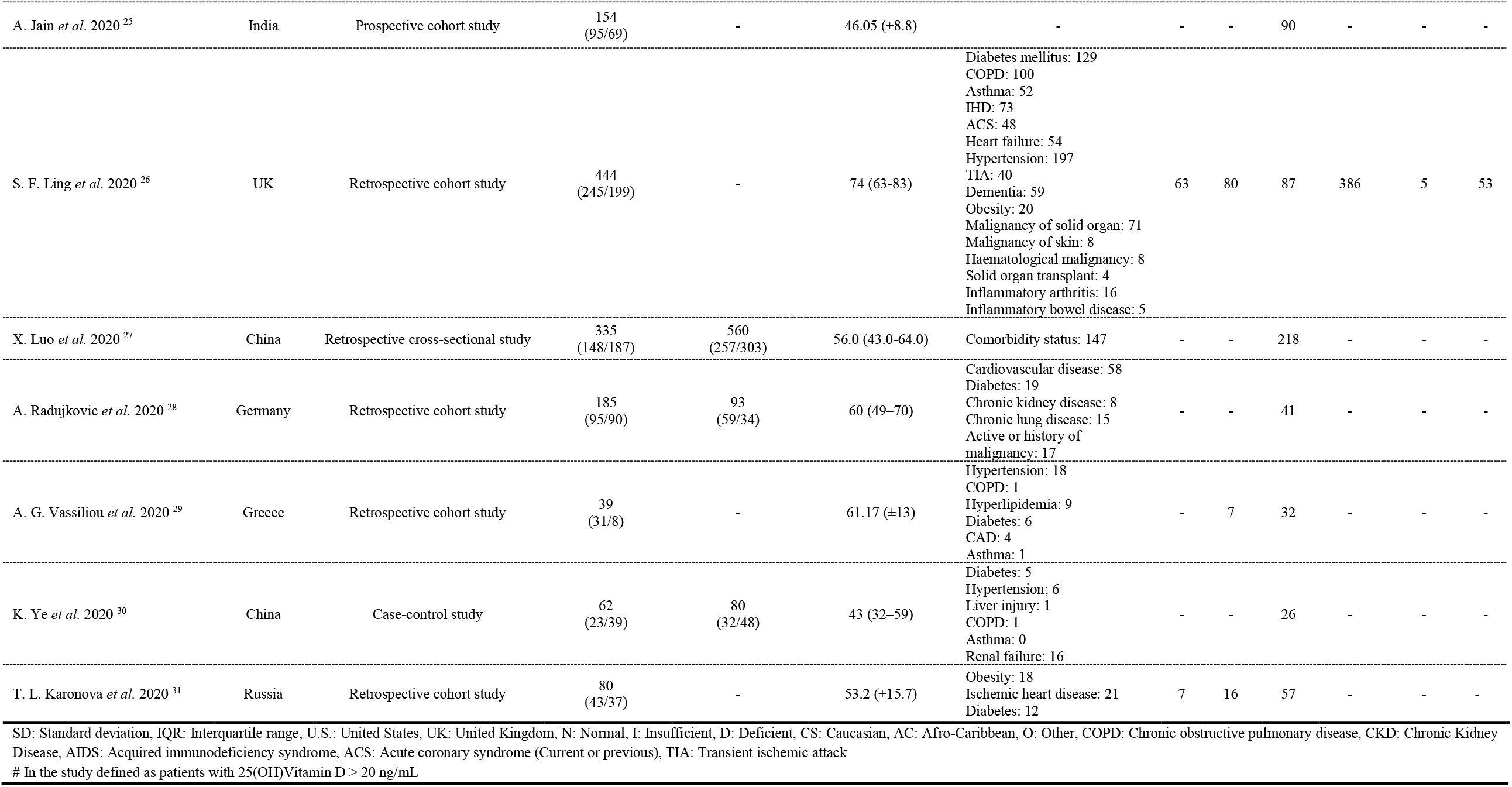
Characteristics of studies entered into the systematic review

| Study | Country | Study design | No. of Patients<br>(Cases)<br>(male/female) | Controls<br>(male/female) | Mean (± SD)<br>Median (IQR) Age<br>of Patients (Cases) | Comorbidity of<br>Patients (Cases) | Vitamin D Status of<br>Patients (Cases) |  |  | Ethnicity of Patients<br>(Cases) |  |  |
| --- | --- | --- | --- | --- | --- | --- | --- | --- | --- | --- | --- | --- |
|  |  |  |  |  |  |  | N | I | D | CS | AC | O |
| J. H. Im *et al.* 2020 <sup>9</sup> | South Korea | Case-control study | 50 | 150 | 57.5 (34.5-68.0) | - | 13 | - | 37 | - | - | - |
| Z. Maghbooli *et al.* 2020 <sup>10</sup> | Iran | Retrospective cross sectional | 235 | - | 58.72 (±15.2)<br>*mean | Diabetes: 86<br>Hypertension: 104<br>Respiratory disease: 72<br>Cancer: 2 | 77 | - | - | - | - | - |
| V. Baktash *et al.* 2020 <sup>11</sup> | UK | Prospective cohort study | 70<br>(42/28) | - | ≥65 | Hypertension: 34<br>Diabetes mellitus: 26<br>Ischemic heart disease: 15<br>Chronic respiratory disease: 13<br>Heart failure: 12<br>Stroke: 9<br>Dementia: 6<br>CKD: 16<br>Atrial fibrillation: 14<br>Cancer: 3<br>Endocrinological disease: 3 | 31 | - | 39 | 50 | - | 20 |
| D. O. Meltzer *et al.* 2020 <sup>12</sup> | U.S. | Retrospective cohort study | 71 | - | - | Hypertension:261<br>Diabetes:137<br>COPD:117<br>Pulmonary circulation disorders: 20<br>Depression :119<br>CKD:116<br>Liver disease :56<br>Comorbidities with immunosuppression: 105 | 39 | - | 32 | - | - | - |
| J. L. Faul *et al.* 2020 <sup>13</sup> | Ireland | Retrospective cross sectional | 33<br>(33/0) | - | ≥40 | - | 21 | - | 12 | 33 | - | - |
| E. Merzon *et al.* 2020 <sup>14</sup> | Israel | Case-control study | 782<br>(385/397) | 7,025<br>(2,849, 4,176) | 35.58 | Depression/Anxiety: 73<br>Schizophrenia: 15<br>Dementia: 27<br>Diabetes mellitus: 154<br>Hypertension: 174<br>Cardiovascular disease: 78<br>Chronic lung disorders: 66<br>Obesity: 235 | 79 | 598 | 105 | - | - | - |
| G. Panagiotou *et al.* 2020 <sup>15</sup> | UK | Retrospective cross sectional | 134<br>(73/61) | - | - | Hypertension: 56<br>Diabetes: 38<br>Obesity: 14<br>Malignancy: 15<br>Respiratory: 42<br>Cardiovascular disease: 20<br>Kidney and Liver diseases: 19 | - | - | 44 | 132 | 1 | 1 |
| G. E. Carpagnano *et al.* 2020 <sup>16</sup> | Italy | Retrospective cohort study | 42<br>(30/12) | - | 65 (±13)<br>*mean | Hypertension: 26<br>Cardiovascular disease: 16<br>CKD: 16<br>Diabetes type II: 11<br>Cerebrovascular disease: 5<br>Psychosis, depression,<br>anxiety: 10<br>Malignancy: 5<br>COPD: 5<br>Asthma: 2 | 8 | 11 | 23 | - | - | - |
| M. D. Nicola *et al.* 2020 <sup>17</sup> | Italy | Retrospective cohort study | 112<br>(52/60) | - | 47.2 (± 16.4) | - | - | - | - | - | - | - |
| F. Macaya *et al.* 2020 <sup>18</sup> | Spain | Retrospective cohort study | 80<br>(35/45) | - | 67.65 (50-84) | Hypertension: 50<br>Diabetes mellitus: 32<br>Cardiac disease: 19 | - | - | 45 | - | - | - |
| S. Karahan *et al.* 2020 <sup>19</sup> | Turkey | Retrospective cohort study | 149<br>(81/68) | - | 63.5 (± 15.3) | Coronary artery disease: 32<br>Hypertension: 85<br>Dyslipidemia: 39<br>Diabetes mellitus: 61<br>Cerebrovascular accident: 9<br>COPD: 15<br>Malignancy: 23<br>CKD: 29<br>Chronic atrial fibrillation: 15<br>Congestive heart failure: 18<br>Acute kidney injury: 16 | 12 | 34 | 103 | - | - | - |
| A. Abdollahi *et al.* 2020 <sup>20</sup> | Iran | Case-control study | 201<br>(66/135) | 201<br>(66/135) | 48 (±16.95) | Hypothyroidism: 15<br>Diabetes mellitus: 42<br>Spleneectomy: 1<br>Heart failure and hypertension: 20<br>Respiratory infections: 14<br>Autoimmune diseases: 11<br>AIDS: 4 | 39 | 161 | 1 | - | - | - |
| C. Arvinte *et al.* 2020 <sup>21</sup> | U.S. | Prospective cohort study<br>(pilot study) | 21<br>(15/6) | - | 60.2 (±17.4)<br>61 (20-94) | - | - | - | - | 4 | - | 17 |
| E. Cereda *et al.* 2020 <sup>22</sup> | Italy | Prospective cohort study | 129<br>(70/59) | - | 77 (65.0-85.0) | COPD: 16<br>Diabetes: 39<br>Hypertension: 89<br>Ischemic heart disease: 52<br>Cancer: 27<br>CKD: 24 | - | 30 # | 99 | - | - | - |
| A. Hamza *et al.* 2020 <sup>23</sup> | Pakistan | Randomized controlled trial study | 168<br>(94/74) | - | 42.26 (±13.69) | - | 22 | 47 | 98 | - | - | - |
| J. L. Hernandez *et al.* 2020 <sup>24</sup> | Spain | Case-control study | 19<br>(7/12) | 197<br>(123/74) | 60.0 (59.0-75.0) | Hypertension: 12<br>Diabetes: 0<br>Cardiovascular disease: 3<br>COPD: 2<br>Active cancer: 0<br>Immunosuppression: 6 | - | - | - | - | - | - |
| A. Jain *et al.* 2020 <sup>25</sup> | India | Prospective cohort study | 154<br>(95/69) | - | 46.05 (±8.8) | - | - | - | 90 | - | - | - |
| S. F. Ling *et al.* 2020 <sup>26</sup> | UK | Retrospective cohort study | 444<br>(245/199) | - | 74 (63-83) | Diabetes mellitus: 129<br>COPD: 100<br>Asthma: 52<br>IHD: 73<br>ACS: 48<br>Heart failure: 54<br>Hypertension: 197<br>TIA: 40<br>Dementia: 59<br>Obesity: 20<br>Malignancy of solid organ: 71<br>Malignancy of skin: 8<br>Haematological malignancy: 8<br>Solid organ transplant: 4<br>Inflammatory arthritis: 16<br>Inflammatory bowel disease: 5 | 63 | 80 | 87 | 386 | 5 | 53 |
| X. Luo *et al.* 2020 <sup>27</sup> | China | Retrospective cross-sectional study | 335<br>(148/187) | 560<br>(257/303) | 56.0 (43.0-64.0) | Comorbidity status: 147 | - | - | 218 | - | - | - |
| A. Radujkovic *et al.* 2020 <sup>28</sup> | Germany | Retrospective cohort study | 185<br>(95/90) | 93<br>(59/34) | 60 (49-70) | Cardiovascular disease: 58<br>Diabetes: 19<br>Chronic kidney disease: 8<br>Chronic lung disease: 15<br>Active or history of malignancy: 17 | - | - | 41 | - | - | - |
| A. G. Vassiliou *et al.* 2020 <sup>29</sup> | Greece | Retrospective cohort study | 39<br>(31/8) | - | 61.17 (±13) | Hypertension: 18<br>COPD: 1<br>Hyperlipidemia: 9<br>Diabetes: 6<br>CAD: 4<br>Asthma: 1 | - | 7 | 32 | - | - | - |
| K. Ye *et al.* 2020 <sup>30</sup> | China | Case-control study | 62<br>(23/39) | 80<br>(32/48) | 43 (32-59) | Diabetes: 5<br>Hypertension: 6<br>Liver injury: 1<br>COPD: 1<br>Asthma: 0<br>Renal failure: 16 | - | - | 26 | - | - | - |
| T. L. Karonova *et al.* 2020 <sup>31</sup> | Russia | Retrospective cohort study | 80<br>(43/37) | - | 53.2 (±15.7) | Obesity: 18<br>Ischemic heart disease: 21<br>Diabetes: 12 | 7 | 16 | 57 | - | - | - |

SD: Standard deviation, IQR: Interquartile range, U.S.: United States, UK: United Kingdom, N: Normal, I: Insufficient, D: Deficient, CS: Caucasian, AC: Afro-Caribbean, O: Other, COPD: Chronic obstructive pulmonary disease, CKD: Chronic Kidney Disease, AIDS: Acquired immunodeficiency syndrome, ACS: Acute coronary syndrome (Current or previous), TIA: Transient ischemic attack

### Quality assessment

Results of quality assessment for studies entered into meta-analysis were fair.

### Publication bias

Begg’s and Egger’s tests findings was as follows for publication bias in main analysis: frequency of vitamin D status (*P*_*B*_=0.38; *P*_*E*_=0.02); mean 25(OH)D concentration (*P*_*B*_=0.80; *P*_*E*_=0.76); vitamin D deficiency and SARS-CoV-2 infection (*P*_*B*_=1.00; *P*_*E*_=0.55); Vitamin D deficiency and COVID-19 severity (*P*_*B*_=0.12; *P*_*E*_=0.14); and vitamin D deficiency and COVID- 19 mortality (*P*_*B*_=0.54; *P*_*E*_=0.62).

### Meta-analysis findings

#### Frequency of Vitamin D status in COVID-19 patients

The meta-analysis of event rates in peer reviewed papers showed that 41% of COVID-19 patients were suffering from vitamin D deficiency (95% CI, 29%-55%), in 42% of patients, levels of vitamin D were lower than the normal range (95% CI, 24%-63%), and only 19% of patients had normal vitamin D levels (95% CI, 11%-32%) (Fig. 2).

**Figure 2.**
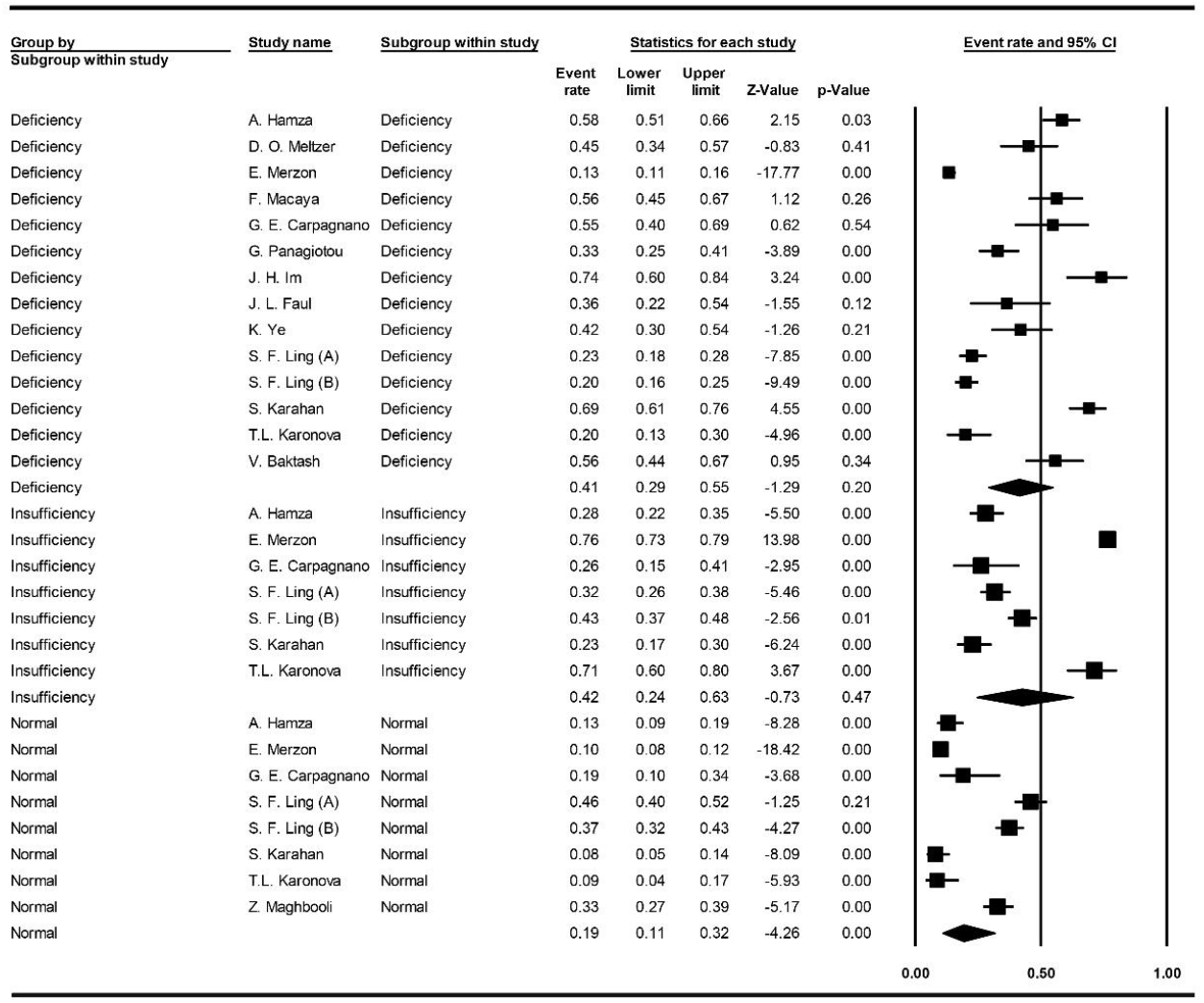
Forest plot for pooling events of vitamin D status.

#### Mean serum 25-hydroxyvitamin D concentration

The meta-analysis of mean 25(OH)D concentration was 20.3 ng/mL among all COVID-19 patients (95% CI, 11.5-28.1), 16.0 ng/mL in severe cases (95% CI, 12.1-19.8) and 24.5 ng/mL in non-severe cases (95% CI, 20.0-29.0) (Fig. 3).

**Figure 3.**
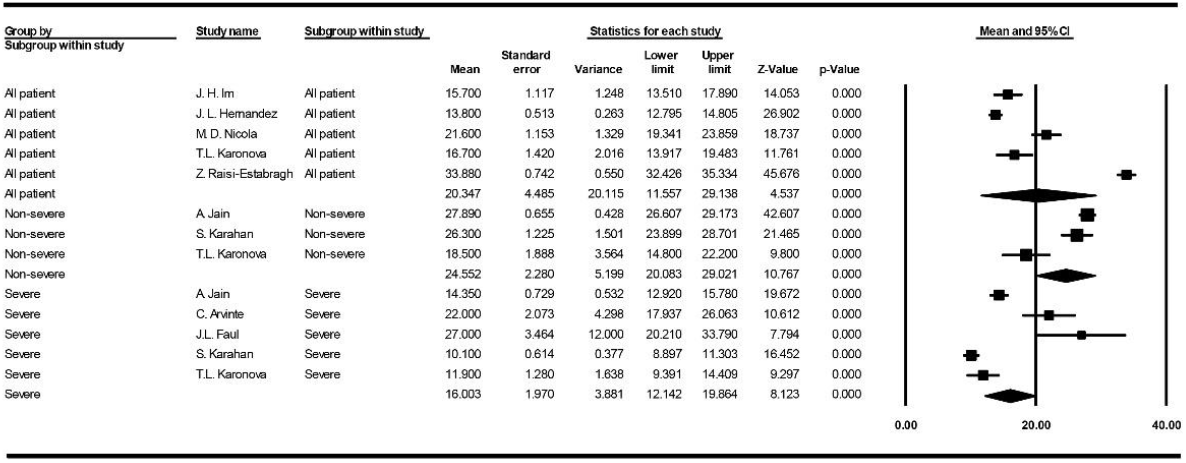
Forest plot for pooling mean 25(OH)D concentrations.

#### Vitamin D Deficiency and SARS-CoV-2 infection

The meta-analysis indicated that odds of getting infected with SARS-CoV-2 increases by 3.3 times in individuals with vitamin D deficiency (95% CI, 2.5-4.3) (Fig. 4).

**Figure 4.**
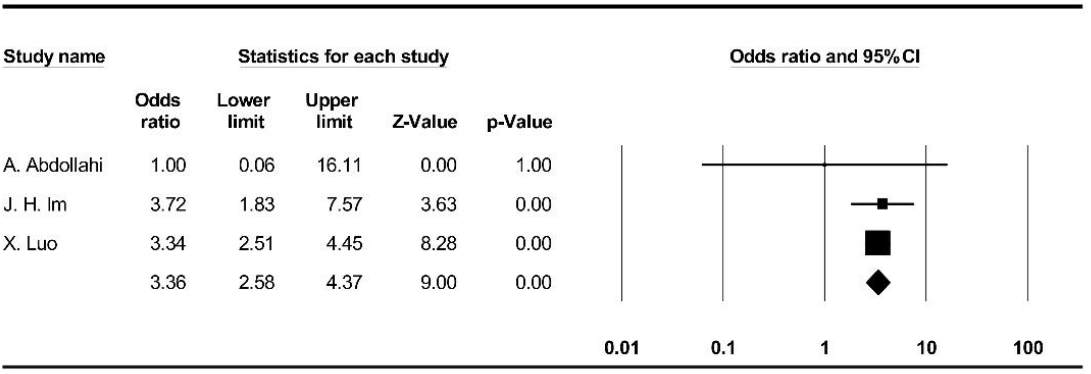
Forest plot for pooling odds ratios of vitamin D deficiency and SARS-CoV-2 infection.

#### Vitamin D Deficiency and COVID-19 severity

The meta-analysis showed that the probability of developing severe stages of COVID-19 is 5.1 times higher in patients with vitamin D deficiency (95% CI, 2.6-10.3) (Fig. 5).

**Figure 5.**
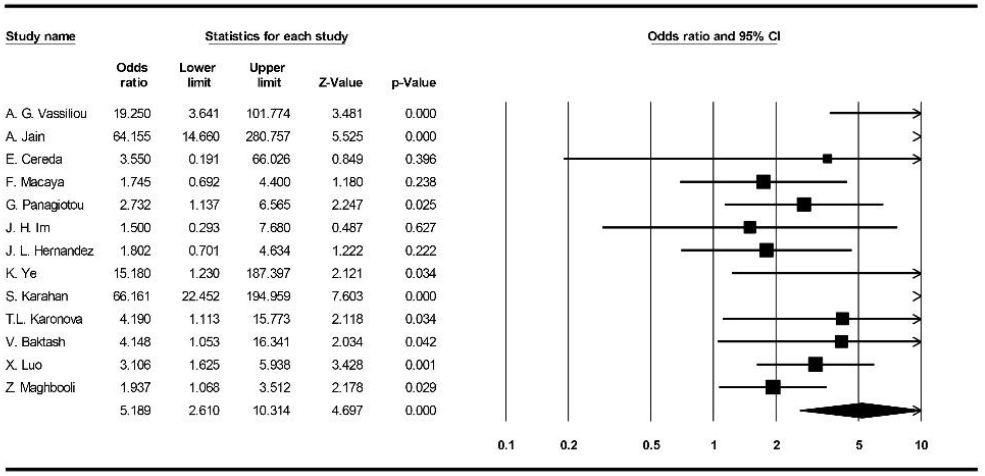
Forest plot for pooling odds ratios of vitamin D deficiency and COVID-19 severity.

#### Vitamin D Deficiency and COVID-19 mortality

The meta-analysis indicated no significant higher COVID-19 mortality related to vitamin D deficient patients (OR: 1.6, 95% CI, 0.5-4.4) (Fig. 6).

**Figure 6.**
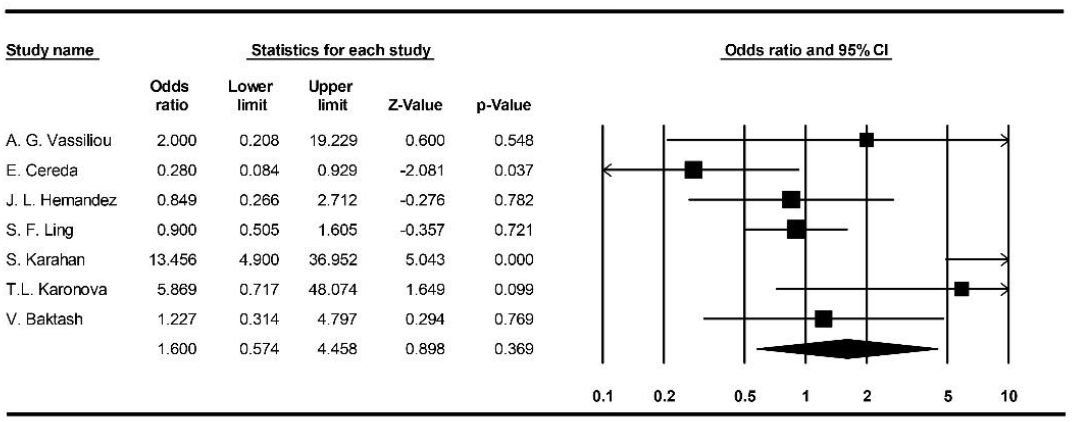
Forest plot for pooling odds ratios of vitamin D deficiency and COVID-19 mortality.

#### Co-morbidities

Meta-analysis of available data on co-morbidities frequency in COVID-19 patients were as follows: in non-severe cases, 13% cancer, 12% chronic kidney disease (CKD), 18% cardiovascular diseases (CVD), 21% diabetes, 29% hypertension (HTN), 12% obesity, and 13% respiratory diseases (Supplementary Figure 1); in severe cases, 13% cancer, 34% CKD, 31% CVD, 35% diabetes, 64% HTsN, 33% obesity, and 17% respiratory diseases (Supplementary Figure 2); in overall, 8% cancer, 20% CKD, 26% CVD, 5% dementia, 15% depression/anxiety, 22% obesity, 26% diabetes, 49% HTN, and 15% respiratory diseases. (Supplementary Figure 3)

#### Ethnicity frequency

Pooling available data regarding ethnicity distribution among COVID-19 patients resulted in 2% Afro-Caribbean, 13% Asian, and 87% Caucasian (Supplementary Figure 4). The results for severe cases were as follows: 2% Asian, 68% Caucasian and 81% Hispanic (Supplementary Figure 5).

## Discussion

### Epidemiological & clinical aspects

Although comparing global statistics of COVID-19 outcomes is difficult, it is clear that the mortality rate is higher in several countries. It seems that among various factors such as age, healthcare system quality, general health status, socioeconomic status, etc. One of the underestimated factors that might be associated with COVID-19 outcome is the vitamin D status in every population. In recent years, vitamin D deficiency/insufficiency has become a global health issue, and its impact has been studied on respiratory viral infections. Most of the epidemiological studies have been reported a higher risk of developing the infection to the severe stages and death in patients with low levels of vitamin D ^32–35^. Besides, vitamin D clinical interventions have demonstrated a significantly reduced risk of respiratory tract infection (RTI), further proposed as a prophylactic or treatment approach against RTIs by WHO in 2017 ^36–38^.

Concerning all of the limitations and lack of high-quality data about the relation of vitamin D status and COVID-19 after several months, we have conducted this systematic review and meta-analysis to maximize the use of every available data, which would give us an overview toward further studies like what we have done recently on the effectiveness of hydroxychloroquine in COVID-19 patients ^39^, which have underestimated first, but the value was revealed after a while.

According to available data entered into our meta-analysis, we could find that approximately 43% of the patients infected with SARS-CoV-2 were suffering from vitamin D deficiency, and this vitamin was insufficient in about 42% of them. We have also found that mean 25(OH)D levels were low (∼20 ng/mL) in all COVID-19 patients. More importantly, our analysis showed that the chance of infecting with SARS-CoV-2 is about three times higher in individuals with vitamin D deficiency and the probability of developing the severe disease in such patients is about 5 times higher than others. However, vitamin D deficiency did not substantially affected mortality rates in such patients.

These findings are in the same line with studies that have debated the association of vitamin D and COVID-19 ^40–44^. Recently, Kaufman *et al*. ^45^, studied the relation of SARS-CoV-2 positivity rates with circulation 25(OH)D among 191,779 patients retrospectively. They found the highest SARS-CoV-2 positivity rate among patients with vitamin D deficiency (12.5%, 95% CI, 12.2-12.8%). Overall, the study indicated a significant inverse relation between SARS-CoV-2 positivity and circulating 25(OH)D levels in COVID-19 patients.

Along with all observational studies, a pilot randomized clinical trial performed by Castillo *et al*. ^46^ on 76 hospitalized COVID-19 patients indicated a promising result for calcifediol therapy in these individuals. In this study, high dose oral calcifediol significantly reduced the need for intensive care unit (ICU) treatment. However, due to the small sample size, more extensive, well-organized clinical trials are needed to robust and confirm this study’s findings.

Additionally, in the case of vitamin D supplements’ benefits against acute respiratory tract infections, Martineau *et al*. conducted a meta-analysis of randomized controlled on 10.933 participants and resulted in an inverse association between vitamin D levels and risk of acute respiratory tract infections. Thus, it can be concluded that patients with lower vitamin D levels or patients with vitamin D deficiency are at higher risk of developing the disease to the severe form ^36^.

### Co-morbidities

After months of investigation on COVID-19, several factors such as male sex, older age, CVD, HTN, chronic lung disease, obesity, and CKD are proposed to be risk factors toward deteriorating COVID-19 patients’ outcomes ^47–50^. Interestingly, one of the conditions that lead to most of the considered risk factors is vitamin D deficiency. Studies indicated that malignancies, diabetes, HTN, and CVDs are significantly related to vitamin D deficiency. Also, studies reported the important role of vitamin D deficiency in older males ^51–53^. Evidence shows that aging, physical activity, obesity, seasonal variation, less vitamin D absorption, pregnancy, thyroid disorders, prolonged use of corticosteroids, ethnicity/race can substantially affect the circulating 25(OH)D levels ^54–60^.

Hence, although studies reported vitamin D deficiency as one of the critical risk factors in clinical outcomes of COVID-19 patients, it seems that it can also be in a strong relationship with basic underlying risk factors and diseases in such patients.

In this case, our analyses indicated that HTN, CVDs, CKDs, diabetes, obesity, and respiratory diseases were the most frequent co-morbidities in COVID-19 patients. According to the facts mentioned above and our findings, it is plausible that both vitamin D deficiency and underlying diseases, which affect each other, may worsen the condition of these patients more than others.

### Ethnicity

From the beginning of the COVID-19 pandemic, different studies have been reported probable associations between COVID-19 and the ethnicity of these patients. Most studies found that the mortality rate among black people is higher than the other ethnic groups ^61–65^. However, other challenges such as human resources, health care systems budgetary, poor management, etc. have to be considered among such people and low-income countries ^66–68^, which unavoidably affects the subject significantly. In recent years, many studies have focused on vitamin D mechanisms and status among various ethnic groups to find the roles of vitamin D and its relationships with any factors or disorders in various ethnicities ^69–72^.

Herein, our findings demonstrated that the most frequent ethnic group has belonged to Caucasians, followed by Hispanic, Asian and Afro-Caribbean. Although there is some evidence on the role of genetic variants in COVID-19 patients, the subject is still not clear enough ^73,74^.

In contrast to many studies about vitamin D status in different ethnicities, Aloia *et al*. have reported that serum 25(OH)D concentration is the same in cross-racial comparison. They found an inconsistency between monoclonal and polyclonal assays for detecting vitamin D binding protein ^75^. Hence, the approach for considering serum 25(OH)D concentration is much important.

### Vitamin D mechanisms & COVID-19

Vitamin D metabolism has been well studied throughout history. Numerous investigations indicate vitamin D’s roles in reducing microbial infections through a physical barrier, natural immunity, and adaptive immunity ^2,76–81^. For example, investigations on respiratory infections indicated that 25(OH)D could effectively induce the host defense peptides against bacterial or viral agents. Vitamin D insufficiency/deficiency can lead to non-communicable and infectious diseases ^2,82,83^. The other potential role of vitamin D is reducing inflammatory induced following SARS-CoV-2 infection by suppressing inflammatory cytokines, reducing leukocytes’ infiltration, interaction with polymorphonuclear leukocytes, and inhibiting complement component C3 ^32,84–88^. Also, according to the available evidence for infections and malignancies ^89,90^, vitamin D may enhance the serological response and CD8^+^ T lymphocytes performance against COVID-19 when the T cells’ exhaustion is related to the critical stages of the disease ^91–93^.

Besides, according to the revealed association of SARS-CoV-2 and angiotensin-converting enzyme 2 (ACE2), this virus can substantially downregulate the ACE2 expression, which seems to leads the COVID-19 patients to deterioration ^94–96^. In contrast, vitamin D affects the renin-angiotensin system pathway and promotes the expression of ACE2 ^97,98^. However, since the high expression of ACE2 can be a risk factor for the severity of the disease ^99^, it is not yet clear enough to conclude how much vitamin D helps the condition. Hence, more evidence and trials are needed to design a treatment plan for three groups of mild, moderate, and severe patients.

It is worth noticing that the current meta-analysis includes the following limitations:1) most of studies entered into the meta-analysis were retrospective in nature; 2) There are inevitable challenges with the reliability of data due to different strategies in a testing (e.g., vitamin D measurement, COVID-19 test, etc.), various subpopulations, etc.; 3) other immunomodulatory factors (e.g., vitamin C, zinc, selenium, etc.), which might be influential in the outcome of COVID-19 patients, have not considered in included studies; and 4) type *II* statistical errors following studies with small sample size. Eventually, to overcome the limitations and bias, the study’s results should be confirmed by robustly large multicentral randomized clinical trials.

## Conclusion

The conditional evidence recommends that vitamin D might be a critical supportive agent for the immune system, mainly in cytokine response regulation against pathogens. In this systematic review and meta-analysis, we found that mean serum 25(OH)D level was low (∼20 ng/mL) in all COVID-19 patients and most of them were suffering from vitamin D deficiency/insufficiency. Also, there is about three times higher chance of getting infected with SARS-CoV-2 among vitamin D deficient individuals and 5 times higher probability of developing the severe disease in such patients. Vitamin D deficiency showed no significant association with mortality rates in these population. The Caucasian was the dominant ethnic group, and the most frequent co-morbidities in COVID-19 patients were HTN, CVDs, CKDs, diabetes, obesity, and respiratory diseases, which might be affected by vitamin D deficiency directly or indirectly. However, further large clinical trials following comprehensive meta-analysis should be taken into account to achieve more reliable findings.

## Data Availability

N/A

## Declarations

### Ethics approval and consent to participate

Not applicable.

### Consent for publication

Not applicable.

### Availability of data and material

Not applicable.

## Acknowledgments

We would like to express our appreciation to the Student Research Committee of Mazandaran University of Medical Sciences for approving this student research proposal with the code 7904. It is also remarkable that the manuscript was published on a pre-print server (available at https://doi.org/10.1101/2020.06.05.20123554).

