## Supplementary for "The Role of Vitamin D in the Age of COVID-19: A Systematic Review and Meta-Analysis"

### *Co-morbidities*

| Group by Subgroup within study | Study name | Subgroup within study | Event rate | Lower limit | Upper limit | Z-Value | p-Value |
|---|---|---|---|---|---|---|---|
| Cancer | G. Panagiotou | Cancer | 0.13 | 0.08 | 0.22 | -6.13 | 0.00 |
| Cancer | S. Karahan | Cancer | 0.13 | 0.06 | 0.26 | -4.40 | 0.00 |
| Cancer | | | 0.13 | 0.08 | 0.20 | -7.54 | 0.00 |
| CKD | F. Macaya | CKD | 0.24 | 0.14 | 0.38 | -3.39 | 0.00 |
| CKD | S. Karahan | CKD | 0.04 | 0.01 | 0.15 | -4.31 | 0.00 |
| CKD | | | 0.12 | 0.02 | 0.48 | -2.04 | 0.04 |
| CVD | F. Macaya | CVD | 0.22 | 0.13 | 0.36 | -3.62 | 0.00 |
| CVD | G. Panagiotou | CVD | 0.16 | 0.10 | 0.25 | -5.80 | 0.00 |
| CVD | S. Karahan | CVD | 0.06 | 0.02 | 0.18 | -4.50 | 0.00 |
| CVD | T.L. Karonova | CVD | 0.24 | 0.14 | 0.37 | -3.69 | 0.00 |
| CVD | | | 0.18 | 0.12 | 0.26 | -6.15 | 0.00 |
| Diabetes | F. Macaya | Diabetes | 0.41 | 0.28 | 0.55 | -1.28 | 0.20 |
| Diabetes | G. Panagiotou | Diabetes | 0.26 | 0.18 | 0.36 | -4.39 | 0.00 |
| Diabetes | K. Ye | Diabetes | 0.06 | 0.02 | 0.17 | -4.62 | 0.00 |
| Diabetes | S. Karahan | Diabetes | 0.26 | 0.15 | 0.40 | -3.20 | 0.00 |
| Diabetes | T.L. Karonova | Diabetes | 0.13 | 0.06 | 0.24 | -4.76 | 0.00 |
| Diabetes | | | 0.21 | 0.12 | 0.34 | -3.99 | 0.00 |
| Hypertension | F. Macaya | Hypertension | 0.41 | 0.28 | 0.55 | -1.28 | 0.20 |
| Hypertension | G. Panagiotou | Hypertension | 0.35 | 0.26 | 0.45 | -2.87 | 0.00 |
| Hypertension | K. Ye | Hypertension | 0.08 | 0.03 | 0.19 | -4.69 | 0.00 |
| Hypertension | S. Karahan | Hypertension | 0.32 | 0.20 | 0.46 | -2.42 | 0.02 |
| Hypertension | | | 0.29 | 0.18 | 0.43 | -2.87 | 0.00 |
| Obesity | F. Macaya | Obesity | 0.27 | 0.16 | 0.41 | -3.15 | 0.00 |
| Obesity | G. Panagiotou | Obesity | 0.05 | 0.02 | 0.12 | -6.21 | 0.00 |
| Obesity | T.L. Karonova | Obesity | 0.11 | 0.05 | 0.22 | -4.86 | 0.00 |
| Obesity | | | 0.12 | 0.05 | 0.30 | -3.50 | 0.00 |
| Respiratory disease | F. Macaya | Respiratory disease | 0.16 | 0.08 | 0.29 | -4.23 | 0.00 |
| Respiratory disease | G. Panagiotou | Respiratory disease | 0.33 | 0.24 | 0.43 | -3.26 | 0.00 |
| Respiratory disease | K. Ye | Respiratory disease | 0.02 | 0.00 | 0.13 | -3.85 | 0.00 |
| Respiratory disease | S. Karahan | Respiratory disease | 0.09 | 0.03 | 0.21 | -4.54 | 0.00 |
| Respiratory disease | | | 0.13 | 0.05 | 0.31 | -3.47 | 0.00 |

Statistics for each study

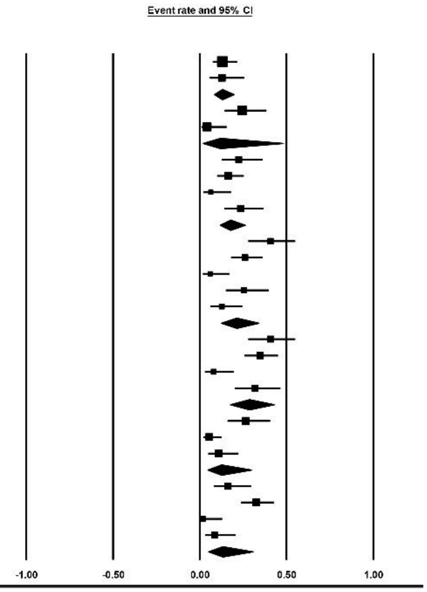


*Supplementary Figure 1. Forest plot for pooling co-morbidities frequency in non-severe cases*

| Group by Subgroup within study | Study name | Subgroup within study | Event rate | Lower limit | Upper limit | Z-Value | p-Value |
|---|---|---|---|---|---|---|---|
| Cancer | G. Panagiotou | Cancer | 0.07 | 0.02 | 0.20 | -4.28 | 0.00 |
| Cancer | S. Karahan | Cancer | 0.17 | 0.11 | 0.25 | -6.06 | 0.00 |
| Cancer | | | 0.13 | 0.06 | 0.26 | -4.27 | 0.00 |
| CKD | F. Macaya | CKD | 0.45 | 0.29 | 0.63 | -0.54 | 0.59 |
| CKD | S. Karahan | CKD | 0.26 | 0.19 | 0.36 | -4.55 | 0.00 |
| CKD | | | 0.34 | 0.19 | 0.54 | -1.60 | 0.11 |
| CVD | F. Macaya | CVD | 0.26 | 0.13 | 0.44 | -2.57 | 0.01 |
| CVD | G. Panagiotou | CVD | 0.12 | 0.05 | 0.26 | -4.20 | 0.00 |
| CVD | S. Karahan | CVD | 0.61 | 0.51 | 0.70 | 2.16 | 0.03 |
| CVD | T.L. Karonova | CVD | 0.32 | 0.17 | 0.52 | -1.76 | 0.08 |
| CVD | | | 0.31 | 0.13 | 0.58 | -1.39 | 0.16 |
| Diabetes | F. Macaya | Diabetes | 0.39 | 0.23 | 0.57 | -1.25 | 0.21 |
| Diabetes | G. Panagiotou | Diabetes | 0.33 | 0.21 | 0.49 | -2.12 | 0.03 |
| Diabetes | K. Ye | Diabetes | 0.20 | 0.05 | 0.54 | -1.75 | 0.08 |
| Diabetes | S. Karahan | Diabetes | 0.48 | 0.39 | 0.58 | -0.40 | 0.69 |
| Diabetes | T.L. Karonova | Diabetes | 0.20 | 0.09 | 0.40 | -2.77 | 0.01 |
| Diabetes | | | 0.35 | 0.25 | 0.47 | -2.44 | 0.01 |
| Hypertension | F. Macaya | Hypertension | 0.97 | 0.80 | 1.00 | 3.35 | 0.00 |
| Hypertension | G. Panagiotou | Hypertension | 0.57 | 0.42 | 0.71 | 0.92 | 0.36 |
| Hypertension | K. Ye | Hypertension | 0.20 | 0.05 | 0.54 | -1.75 | 0.08 |
| Hypertension | S. Karahan | Hypertension | 0.69 | 0.59 | 0.77 | 3.67 | 0.00 |
| Hypertension | | | 0.64 | 0.40 | 0.83 | 1.17 | 0.24 |
| Obesity | F. Macaya | Obesity | 0.32 | 0.18 | 0.50 | -1.93 | 0.05 |
| Obesity | G. Panagiotou | Obesity | 0.21 | 0.12 | 0.36 | -3.46 | 0.00 |
| Obesity | T.L. Karonova | Obesity | 0.48 | 0.30 | 0.67 | -0.20 | 0.84 |
| Obesity | | | 0.33 | 0.20 | 0.49 | -2.05 | 0.04 |
| Respiratory disease | F. Macaya | Respiratory disease | 0.16 | 0.07 | 0.33 | -3.38 | 0.00 |
| Respiratory disease | G. Panagiotou | Respiratory disease | 0.29 | 0.17 | 0.44 | -2.68 | 0.01 |
| Respiratory disease | S. Karahan | Respiratory disease | 0.11 | 0.06 | 0.18 | -6.62 | 0.00 |
| Respiratory disease | | | 0.17 | 0.09 | 0.31 | -3.96 | 0.00 |

Statistics for each study

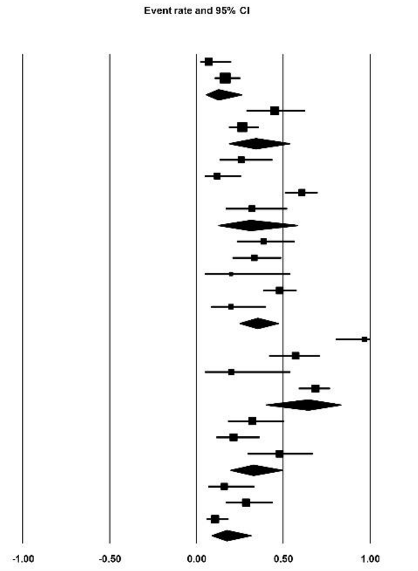


*Supplementary Figure 2. Forest plot for pooling co-morbidities frequency in severe cases*

| Group by Subgroup within study | Study name | Subgroup within study | Statistics for each study: Event rate | Lower limit | Upper limit | Z-Value | p-Value |
|---|---|---|---|---|---|---|---|
| Cancer | A. Radujkovic | Cancer | 0.09 | 0.06 | 0.14 | -9.00 | 0.00 |
| Cancer | E. Cereda | Cancer | 0.21 | 0.15 | 0.29 | -6.14 | 0.00 |
| Cancer | G. E. Carpagnano | Cancer | 0.12 | 0.05 | 0.26 | -4.20 | 0.00 |
| Cancer | G. Panagiotou | Cancer | 0.11 | 0.07 | 0.18 | -7.56 | 0.00 |
| Cancer | J. L. Hernandez | Cancer | 0.04 | 0.02 | 0.07 | -8.58 | 0.00 |
| Cancer | S. Karahan | Cancer | 0.15 | 0.10 | 0.22 | -7.50 | 0.00 |
| Cancer | V. Baktash | Cancer | 0.04 | 0.01 | 0.12 | -5.26 | 0.00 |
| Cancer | Z. Maghbooli | Cancer | 0.01 | 0.00 | 0.03 | -6.70 | 0.00 |
| Cancer | | | 0.08 | 0.05 | 0.14 | -8.35 | 0.00 |
| CKD | A. Radujkovic | CKD | 0.04 | 0.02 | 0.08 | -8.57 | 0.00 |
| CKD | E. Cereda | CKD | 0.19 | 0.13 | 0.26 | -6.52 | 0.00 |
| CKD | F. Macaya | CKD | 0.33 | 0.23 | 0.43 | -3.06 | 0.00 |
| CKD | G. E. Carpagnano | CKD | 0.38 | 0.25 | 0.53 | -1.53 | 0.13 |
| CKD | S. Karahan | CKD | 0.19 | 0.14 | 0.27 | -6.86 | 0.00 |
| CKD | V. Baktash | CKD | 0.23 | 0.15 | 0.34 | -4.27 | 0.00 |
| CKD | | | 0.20 | 0.12 | 0.31 | -4.67 | 0.00 |
| CVD | A. Abdollahi | CVD | 0.10 | 0.07 | 0.15 | -9.35 | 0.00 |
| CVD | A. Radujkovic | CVD | 0.31 | 0.25 | 0.38 | -4.95 | 0.00 |
| CVD | E. Cereda | CVD | 0.40 | 0.32 | 0.49 | -2.19 | 0.03 |
| CVD | E. Merzon | CVD | 0.10 | 0.08 | 0.12 | -18.44 | 0.00 |
| CVD | F. Macaya | CVD | 0.24 | 0.16 | 0.34 | -4.44 | 0.00 |
| CVD | G. E. Carpagnano | CVD | 0.38 | 0.25 | 0.53 | -1.53 | 0.13 |
| CVD | G. Panagiotou | CVD | 0.15 | 0.10 | 0.22 | -7.18 | 0.00 |
| CVD | J. L. Hernandez | CVD | 0.11 | 0.07 | 0.16 | -9.21 | 0.00 |
| CVD | S. Karahan | CVD | 0.44 | 0.36 | 0.52 | -1.55 | 0.12 |
| CVD | T.L. Karonova | CVD | 0.26 | 0.18 | 0.37 | -4.07 | 0.00 |
| CVD | V. Baktash | CVD | 0.71 | 0.60 | 0.81 | 3.46 | 0.00 |
| CVD | | | 0.26 | 0.16 | 0.39 | -3.53 | 0.00 |
| Dementia | E. Merzon | Dementia | 0.03 | 0.02 | 0.05 | -17.01 | 0.00 |
| Dementia | V. Baktash | Dementia | 0.09 | 0.04 | 0.18 | -5.54 | 0.00 |
| Dementia | | | 0.05 | 0.02 | 0.12 | -6.14 | 0.00 |
| Depression/Anxiety | E. Merzon | Depression/Anxiety | 0.09 | 0.07 | 0.12 | -18.50 | 0.00 |
| Depression/Anxiety | G. E. Carpagnano | Depression/Anxiety | 0.24 | 0.13 | 0.39 | -3.21 | 0.00 |
| Depression/Anxiety | | | 0.15 | 0.05 | 0.33 | -3.20 | 0.00 |
| Obesity | E. Merzon | Obesity | 0.30 | 0.27 | 0.33 | -10.83 | 0.00 |
| Obesity | F. Macaya | Obesity | 0.29 | 0.20 | 0.40 | -3.67 | 0.00 |
| Obesity | G. Panagiotou | Obesity | 0.10 | 0.06 | 0.17 | -7.61 | 0.00 |
| Obesity | T.L. Karonova | Obesity | 0.22 | 0.15 | 0.33 | -4.62 | 0.00 |
| Obesity | | | 0.22 | 0.14 | 0.33 | -4.60 | 0.00 |
| Diabetes | A. Abdollahi | Diabetes | 0.21 | 0.16 | 0.27 | -7.67 | 0.00 |
| Diabetes | A. Radujkovic | Diabetes | 0.10 | 0.07 | 0.16 | -8.95 | 0.00 |
| Diabetes | E. Cereda | Diabetes | 0.30 | 0.23 | 0.39 | -4.36 | 0.00 |
| Diabetes | E. Merzon | Diabetes | 0.20 | 0.17 | 0.23 | -15.63 | 0.00 |
| Diabetes | F. Macaya | Diabetes | 0.40 | 0.30 | 0.51 | -1.78 | 0.08 |
| Diabetes | G. E. Carpagnano | Diabetes | 0.26 | 0.15 | 0.41 | -2.95 | 0.00 |
| Diabetes | G. Panagiotou | Diabetes | 0.28 | 0.21 | 0.37 | -4.84 | 0.00 |
| Diabetes | J. L. Hernandez | Diabetes | 0.17 | 0.13 | 0.23 | -8.31 | 0.00 |
| Diabetes | S. Karahan | Diabetes | 0.41 | 0.33 | 0.49 | -2.20 | 0.03 |
| Diabetes | T.L. Karonova | Diabetes | 0.15 | 0.09 | 0.25 | -5.54 | 0.00 |
| Diabetes | V. Baktash | Diabetes | 0.37 | 0.27 | 0.49 | -2.13 | 0.03 |
| Diabetes | Z. Maghbooli | Diabetes | 0.37 | 0.31 | 0.43 | -4.06 | 0.00 |
| Diabetes | | | 0.26 | 0.20 | 0.32 | -6.76 | 0.00 |
| Hypertension | A. G. Vassiliou | Hypertension | 0.46 | 0.31 | 0.62 | -0.48 | 0.63 |
| Hypertension | E. Cereda | Hypertension | 0.69 | 0.61 | 0.76 | 4.20 | 0.00 |
| Hypertension | E. Merzon | Hypertension | 0.22 | 0.19 | 0.25 | -14.55 | 0.00 |
| Hypertension | F. Macaya | Hypertension | 0.63 | 0.51 | 0.72 | 2.21 | 0.03 |
| Hypertension | G. E. Carpagnano | Hypertension | 0.62 | 0.47 | 0.75 | 1.53 | 0.13 |
| Hypertension | G. Panagiotou | Hypertension | 0.42 | 0.34 | 0.50 | -1.89 | 0.06 |
| Hypertension | J. L. Hernandez | Hypertension | 0.39 | 0.32 | 0.46 | -3.18 | 0.00 |
| Hypertension | S. Karahan | Hypertension | 0.57 | 0.49 | 0.65 | 1.71 | 0.09 |
| Hypertension | V. Baktash | Hypertension | 0.49 | 0.37 | 0.60 | -0.24 | 0.81 |
| Hypertension | Z. Maghbooli | Hypertension | 0.44 | 0.38 | 0.51 | -1.76 | 0.08 |
| Hypertension | | | 0.49 | 0.37 | 0.60 | -0.23 | 0.82 |
| Respiratory disease | A. Radujkovic | Respiratory disease | 0.08 | 0.05 | 0.13 | -9.01 | 0.00 |
| Respiratory disease | E. Cereda | Respiratory disease | 0.12 | 0.08 | 0.19 | -7.32 | 0.00 |
| Respiratory disease | E. Merzon | Respiratory disease | 0.08 | 0.07 | 0.11 | -18.53 | 0.00 |
| Respiratory disease | F. Macaya | Respiratory disease | 0.16 | 0.10 | 0.26 | -5.41 | 0.00 |
| Respiratory disease | G. E. Carpagnano | Respiratory disease | 0.17 | 0.08 | 0.31 | -3.89 | 0.00 |
| Respiratory disease | G. Panagiotou | Respiratory disease | 0.31 | 0.24 | 0.40 | -4.21 | 0.00 |
| Respiratory disease | J. L. Hernandez | Respiratory disease | 0.08 | 0.05 | 0.12 | -9.29 | 0.00 |
| Respiratory disease | S. Karahan | Respiratory disease | 0.10 | 0.06 | 0.16 | -8.04 | 0.00 |
| Respiratory disease | V. Baktash | Respiratory disease | 0.19 | 0.11 | 0.29 | -4.81 | 0.00 |
| Respiratory disease | Z. Maghbooli | Respiratory disease | 0.31 | 0.25 | 0.37 | -5.77 | 0.00 |
| Respiratory disease | | | 0.15 | 0.09 | 0.22 | -7.09 | 0.00 |

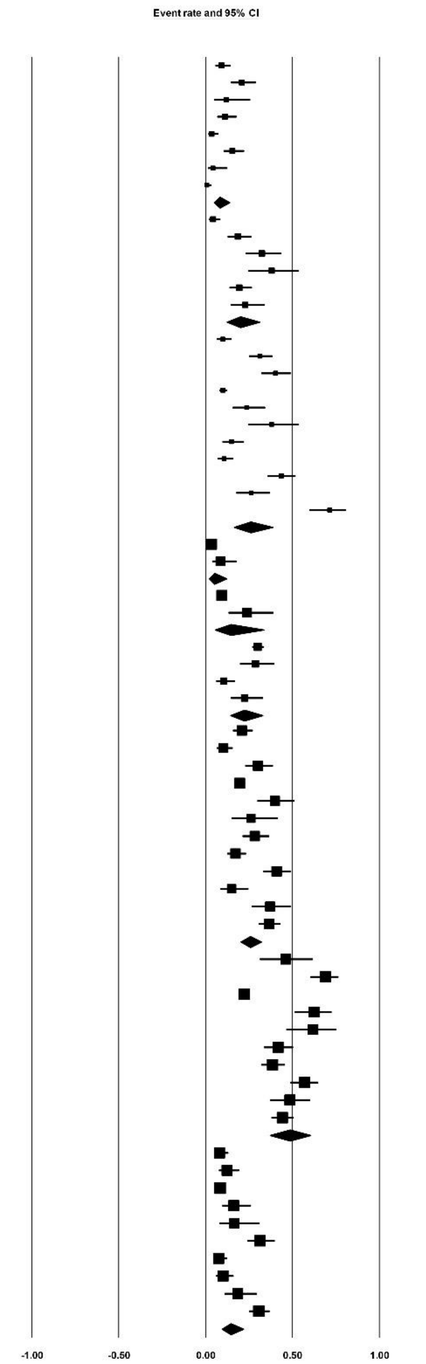


*Supplementary Figure 3. Forest plot for pooling overall co-morbidities frequency in all studies*

| Group by Subgroup within study | Study name | Subgroup within study | Event rate | Lower limit | Upper limit | Z-Value | p-Value |
|---|---|---|---|---|---|---|---|
| Afro-Caribbean | G. Panagiotou | Afro-Caribbean | 0.01 | 0.00 | 0.05 | -4.87 | 0.00 |
| Afro-Caribbean | S. F. Ling (A) | Afro-Caribbean | 0.01 | 0.00 | 0.03 | -6.68 | 0.00 |
| Afro-Caribbean | S. F. Ling (B) | Afro-Caribbean | 0.04 | 0.02 | 0.07 | -10.90 | 0.00 |
| Afro-Caribbean | V. Baktash | Afro-Caribbean | 0.01 | 0.00 | 0.09 | -4.20 | 0.00 |
| Afro-Caribbean | | | 0.02 | 0.01 | 0.05 | -8.01 | 0.00 |
| Asian | G. Panagiotou | Asian | 0.03 | 0.01 | 0.08 | -6.86 | 0.00 |
| Asian | S. F. Ling (A) | Asian | 0.06 | 0.04 | 0.10 | -9.99 | 0.00 |
| Asian | S. F. Ling (B) | Asian | 0.28 | 0.23 | 0.33 | -7.61 | 0.00 |
| Asian | V. Baktash | Asian | 0.29 | 0.19 | 0.40 | -3.46 | 0.00 |
| Asian | | | 0.13 | 0.05 | 0.29 | -3.65 | 0.00 |
| Caucasian | G. Panagiotou | Caucasian | 0.96 | 0.90 | 0.98 | 7.33 | 0.00 |
| Caucasian | J. L. Faul | Caucasian | 0.99 | 0.78 | 1.00 | 2.77 | 0.01 |
| Caucasian | S. F. Ling (A) | Caucasian | 0.92 | 0.88 | 0.95 | 10.07 | 0.00 |
| Caucasian | S. F. Ling (B) | Caucasian | 0.61 | 0.55 | 0.66 | 3.78 | 0.00 |
| Caucasian | V. Baktash | Caucasian | 0.71 | 0.60 | 0.81 | 3.46 | 0.00 |
| Caucasian | | | 0.87 | 0.68 | 0.96 | 3.25 | 0.00 |

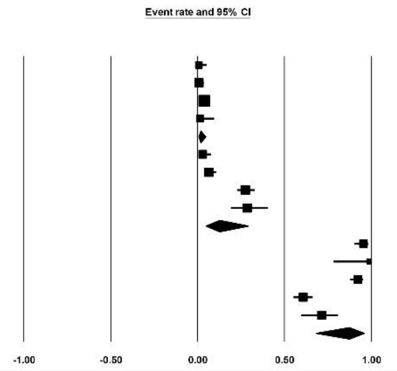


*Supplementary Figure 4. Forest plot for pooling overall ethnicity frequency in all studies*

| Group by Subgroup within study | Study name | Subgroup within study | Event rate | Lower limit | Upper limit | Z-Value | p-Value |
|---|---|---|---|---|---|---|---|
| Asian | G. Panagiotou | Asian | 0.02 | 0.00 | 0.15 | -3.67 | 0.00 |
| Asian | | | 0.02 | 0.00 | 0.15 | -3.67 | 0.00 |
| Caucasian | C. Arvinte | Caucasian | 0.19 | 0.07 | 0.41 | -2.60 | 0.01 |
| Caucasian | G. Panagiotou | Caucasian | 0.95 | 0.83 | 0.99 | 4.13 | 0.00 |
| Caucasian | | | 0.68 | 0.03 | 0.99 | 0.34 | 0.74 |
| Hispanic | C. Arvinte | Hispanic | 0.81 | 0.59 | 0.93 | 2.60 | 0.01 |
| Hispanic | | | 0.81 | 0.59 | 0.93 | 2.60 | 0.01 |

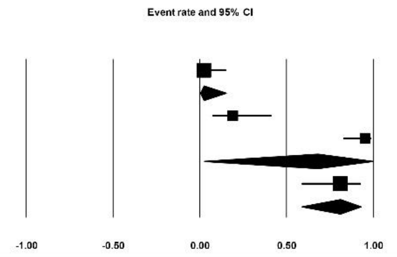


*Supplementary Figure 5. Forest plot for pooling ethnicity frequency in severe cases*
